# Deep Learning Segmentation of Glomeruli on Kidney Donor Frozen Sections

**DOI:** 10.1101/2021.09.16.21263707

**Authors:** Richard C. Davis, Xiang Li, Yuemei Xu, Zehan Wang, Nao Souma, Gina Sotolongo, Jonathan Bell, Matthew Ellis, David Howell, Xiling Shen, Kyle Lafata, Laura Barisoni

## Abstract

**Purpose:** Recent advances in computational image analysis offer the opportunity to develop automatic quantification of histologic parameters as aid tools for practicing pathologists. This work aims to develop deep learning (DL) models to quantify non-sclerotic and sclerotic glomeruli on frozen sections from donor kidney biopsies.

**Approach:** A total of 258 whole slide images (WSI) from cadaveric donor kidney biopsies performed at our institution (n=123) and at external institutions (n=135) were used in this study. WSIs from our institution were divided at the patient level into training and validation datasets (Ratio: 0.8:0.2) and external WSIs were used as an independent testing dataset. Non-sclerotic (n=22767) and sclerotic (n=1366) glomeruli were manually annotated by study pathologists on all WSIs. A 9-layer convolutional neural network based on the common U-Net architecture was developed and tested for the segmentation of non-sclerotic and sclerotic glomeruli. DL-derived, manual segmentation and reported glomerular count (standard of care) were compared.

**Results:** The average Dice Similarity Coefficient testing was 0.90 and 0.83. and the F1, Recall, and Precision scores were 0.93, 0.96, and 0.90, and 0.87, 0.93, and 0.81, for non-sclerotic and sclerotic glomeruli, respectively. DL-derived and manual segmentation derived glomerular counts were comparable, but statistically different from reported glomerular count.

**Conclusions:** DL segmentation is a feasible and robust approach for automatic quantification of glomeruli. This work represents the first step toward new protocols for the evaluation of donor kidney biopsies.

## 1. Introduction

Renal allograft transplantation has superior long-term survival compared to dialysis^1^. However, fewer allografts remain available for transplantation than the number of patients on the transplant waiting list^2–4^. To address this problem, the extended criteria donor (ECD) program was introduced in 2002 in the United States, which allowed for transplantation of allografts from cadaveric donors > 60 years of age and those with comorbidities^2^. To increase the utilization of cadaveric kidneys and improve the prediction of their function, the ECD was later substituted by the Kidney Donor Profile Index (KDPI)^5,6^. While the utility of morphologic parameters to predict allograft function and outcomes is controversial ^4,5,7–13^, histologic analyses of frozen sections of kidney wedge biopsies stained with hematoxylin and eosin (H&E) remain the current practice in North America for determining the suitability of cadaveric kidneys for transplantation.

Semi-quantitative assessment of interstitial fibrosis and inflammation, tubular atrophy and acute tubular injury, arterial intimal fibrosis and arteriolar hyalinosis, presence of intravascular thrombi or glomerular pathology, and percent of globally sclerotic glomeruli are routinely evaluated prior to implantation to prognosticate post-transplant kidney function^14^. The overall longevity of the allograft, however, depends also on other post-transplant superimposed events, such as episodes of T cell or antibody mediated rejection, measured by the Banff scores^15^, response to immunosuppression, viral infections, and other recipient-specific clinical conditions.

More recently, the i-Box, integrating demographic, clinical and morphologic data, was proven to be a useful tool to better predict allograft long term outcomes^16^. Semi-quantitative features, such as interstitial fibrosis, tubular atrophy, arteriosclerosis (i.e. arterial intimal fibrosis), arteriolar hyalinosis, interstitial inflammation, and percent of globally sclerotic glomeruli are routinely evaluated prior to implantation, often by pathologists on-call who do not always have a kidney domain expertise^3–5,8,9^, resulting in implantation of suboptimal donor renal allografts or improper discarding of otherwise suitable allografts^3,5^. High inter- and intra-observer variability further complicate this paradigm ^14,17^.

Since the introduction of commercial whole slide scanners in 1998, digital pathology has become an increasingly important aspect of pathology workflows in both research and clinical practice ^18–22^. Digital pathology enables computational image analysis to be applied to digitized tissue samples. In particular, deep learning (DL), a specific type of machine learning ^23^, is a useful tool for image representation and image analysis tasks^24^. DL methods have been implemented for a wide array of digital pathology domains^25,26^, such as cell detection^27^ and segmentation^28^, detection of breast cancer metastases in lymph nodes^29^, and grading of gliomas^30^. DL approaches have also been applied to kidney biopsies for the automatic detection of normal structures (e.g., glomeruli, urinary space, tubules, and vessels)^28^, and abnormal structures (e.g., global sclerosis, interstitial fibrosis and tubular atrophy)^14,31–36^, using WSIs derived from formalin-fixed and paraffin embedded sections. However, DL studied on frozen sections of kidney remain limited^37^.

Here, we present a DL approach to automatically detect and segment sclerotic and non-sclerotic glomeruli on frozen sections of donor kidney biopsies. We anticipate that this initial pipeline can be enriched by the DL segmentation and quantification of other relevant histologic parameters, resulting into a robust interactive human-machine protocol for the assessment of donor kidney biopsies with potential scalability in clinical practice.

## 2. Materials and Methods

### 2.1 Whole Slide Imaging Dataset

This study was approved by the Duke University Institutional Review Board. A total 211 kidney donors deceased between January 2015 and January 2020 were included in the study, for a total of 268 frozen section H&E-stained slides. Of these, 75 donors had a wedge kidney biopsy performed, frozen, cut, and stained with Hematoxylin & Eosin (H&E) at Duke University Medical Center (DUMC) for a total of 128 frozen section H&E-stained glass slides (Internal cases). Of these 75 donors, 53 had bilateral biopsies and 22 had unilateral biopsies performed. The remaining 136 deceased donors had a wedge kidney biopsy performed, frozen, cut, and stained with H&E in other institutions (External cases) and subsequently reviewed at DUMC, for a total of 140 frozen section H&E-stained slides (Figure 1 - a1).

**Figure 1:**
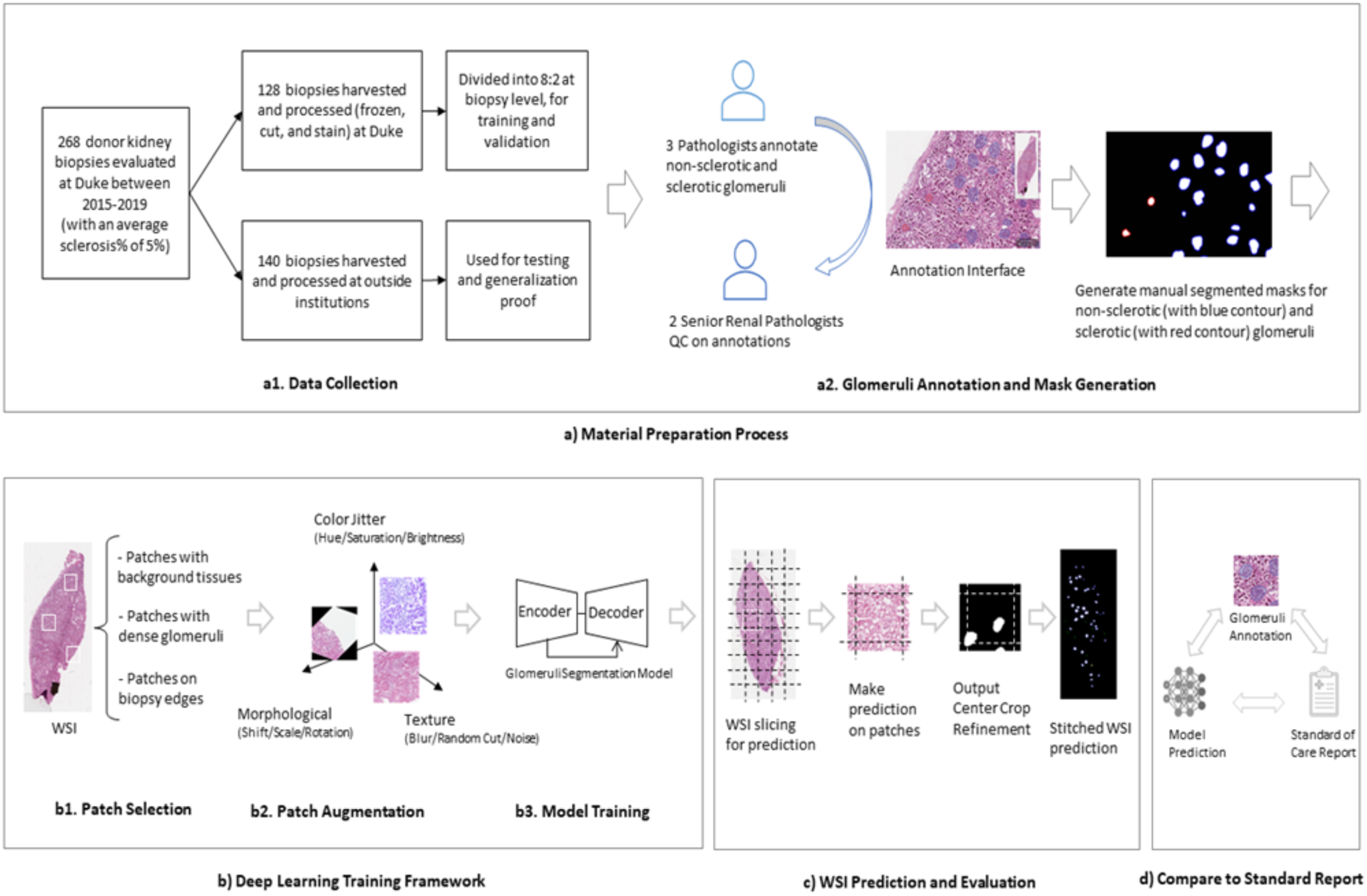
Overall research design. **(a)** The material preparation process consisted of collection of cases from Duke University medical center and outside institutions, followed by annotation by 3 pathologists, QC by expert renal pathologists and mask generation. **(b)** The training process includes selection and augmentation of training samples, followed by model optimization for segmenting the glomeruli. **(c)** Predictions are made based on sequential patches, which are stitched together to recover a WSI prediction. **(d)** The model performance is further investigated by comparing to the standard of care report.

Whole slide images (WSIs) were acquired at 40X magnification on a Leica AT2 whole-slide scanner located in the Duke University Department of Pathology’s BioRepository and Precision Pathology Center for all cases. All WSI were reviewed for image quality assurance, and 10 WSI excluded because they had with severe artifacts, including excessive folding, poor quality of staining, and the presence of bubbles under the coverslip. The final WSI dataset included a total of 258 WSIs (123 Internal and 135 External).

### 2.2 Manual Segmentation of Glomeruli

Three primary annotators, 1 post-graduate year one (GS) and 1 post-graduate year two (RD) trainees in the Duke accredited residency program, and 1 internationally trained pathologist (YM), manually segmented non-sclerotic glomeruli and globally sclerotic glomeruli. Segmentation was achieved by manually outlining glomeruli in all 258 WSIs using a publicly available digital pathology tool (QuPath, version 0.1.2) ^8,13^. For non-sclerotic glomeruli, annotations were made by tracing the Bowman’s capsule and through the vascular and tubular pole of the glomerulus, when visible, to maintain a continuous circular annotation outline of the individual glomeruli. As the Bowman’s capsule of globally sclerotic glomeruli is generally inconsistent or separated from the tuft by a white space representing a processing/freezing artifact, only the sclerosed tuft was outlined during the annotation process (Figure 2).

**Figure 2:**
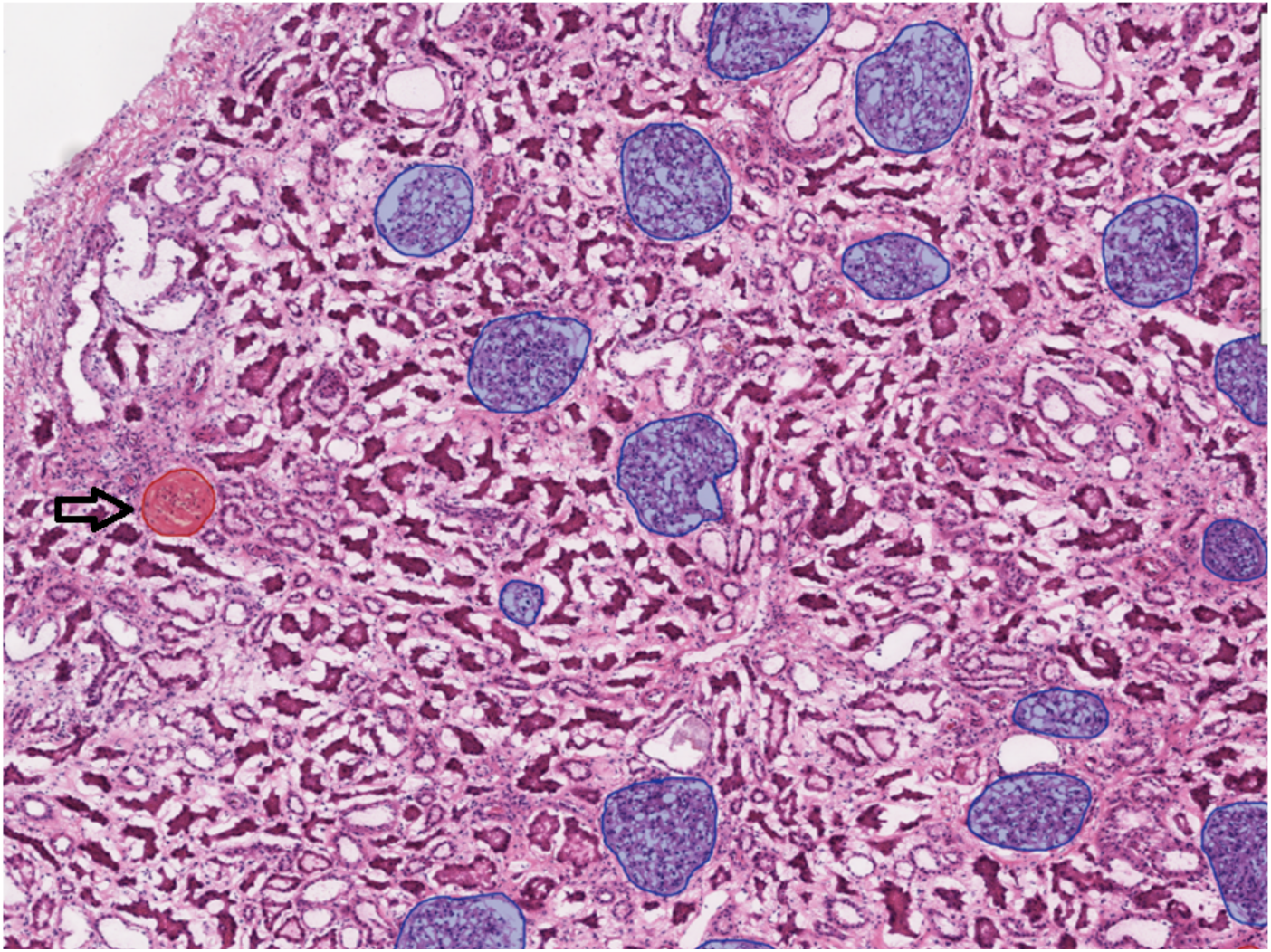
An illustrating example of a whole slide image with glomerular segmentations. Manual segmentations of non-sclerotic glomeruli (blue circle) and sclerotic glomeruli (red circle – black arrow) on whole slide images are generated using QuPath.

All manual segmentations were reviewed by two expert renal pathologists (LB & DH) to assure accuracy of glomerular detection and boundaries. The expert pathologists traced non-sclerotic and sclerotic glomeruli when missed by the primary annotators and retraced the boundaries in those glomeruli where the primary annotator did not follow the segmentation criteria. Matched pairs of shift-invariant WSIs and manual segmentations were considered as DL training samples (Figure 1 - a2).

### 2.3 Deep Learning Implementation and Performance Evaluation

The 75 Internal cases (123 WSIs) were divided at the patient level into training (D1) and validation (D2) datasets with an 0.8:0.2 ratio, respectively, and used to train/fine-tune the DL model. The 135 External cases (135 WSIs) were used as an independent testing dataset (D3).

#### 2.3.1 Network architecture

A DL framework was developed to automatically identify and segment non-sclerotic vs. globally sclerotic glomeruli on frozen section WSIs. A 9-module convolutional neural network (CNN) based on the common U-Net architecture^38^ with a dilated bottleneck was developed for glomerular segmentation (Figure 3). Our U-Net architecture is a symmetric encoder-decoder fully convolutional network with a 256×256×3 input layer that produces pixel-level segmentation results. Each encoder module contains two convolution blocks consisting of a 3×3 convolutional layer, a batch normalization procedure, and a rectified linear unit (ReLU) activation function. Modules are connected by a 2×2 max-pooling layer for down-sampling.

**Figure 3:**
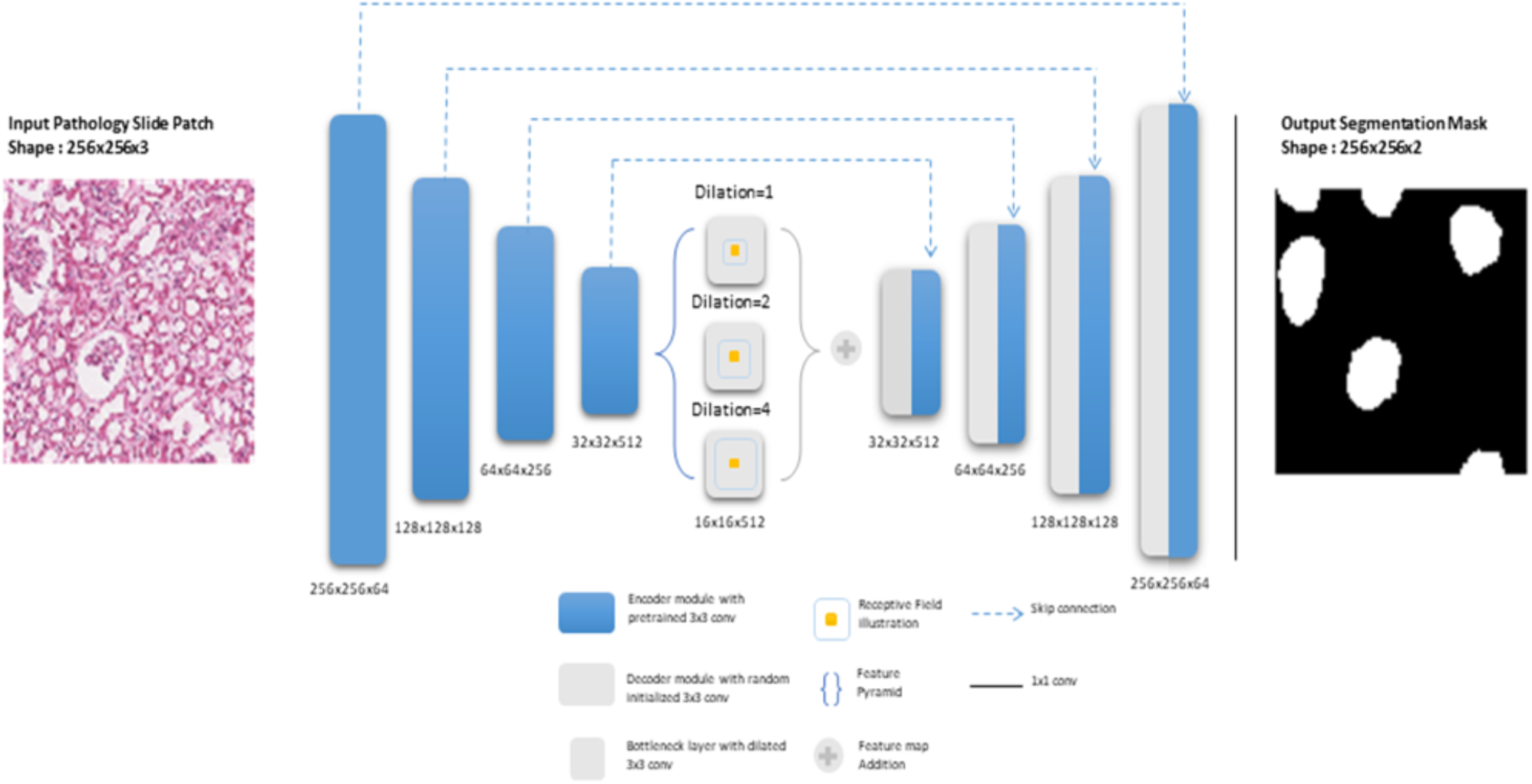
Deep learning model architecture. An input glomeruli patch (left) of size 256×256×3 is fed into a 9-module U-Net model. For this model, each of the four encoder (i.e., left side of the network) modules are loaded with VGG pre-trained weights, which are then fine-tuned during training. The bottleneck module consists of three dilated convolution layers with different dilation rates. The feature maps at this level are added element-wise at the end of the bottleneck. The four decoder modules (i.e., right side of the network) use transpose convolutions to up-sample the feature maps, and skip-connections incorporate the encoder information. Finally, a 1×1 convolution layer maps the information to a 256×256 binary image of pixel-level predictions of glomerular locations.

The down-sampled bottleneck module consists of three dilation operations^39^ for each convolutional layer, which enlarge the receptive field to capture coarse field-of-view imaging details within the high-level feature maps. The decoder modules recover and up-sample the semantic information generated from the bottleneck module, each of which includes a 2×2 transpose convolution block and a 3×3 convolution block. Long-term skip connections are used for enhancing different scale texture details provided in the encoding layers. Finally, a 1×1 convolutional layer is used to output a 2-layer probability map representing glomerular foreground vs. background.

#### 2.3.2 Model training and validation

Two independent models were trained in parallel for normal and globally sclerotic glomeruli, respectively. Using the training data, random 256×256 image patches were extracted at 5x magnification from regions of relatively high glomerular density. Matched pairs of shift-invariant image patches and corresponding manual segmentations were used as training examples to train the DL model. To boost model generalization, a data augmentation procedure was applied, including basic operations (e.g., Horizontal Vertical Flip, Crop Resize), morphological transformations (e.g., Shift Scale Rotate, Elastic Transform), color distortions (e.g., Contrast Brightness, Hue Saturation Value), and other image processing operations (Gaussian Blur, Random Cutout).

Training utilized transfer learning, where the network’s first four fully convolutional blocks were initialized as the pre-trained ImageNet^40^ weights of the publicly available VGG16^31^ data. During the training process, a cross entropy loss function was minimized based on Adam optimization^32^ to learn optimal model hyper-parameterization. Training was run for 200 epochs with a batch size of 16 input patches and an initial learning rate of 0.001. The training implementation achieving the lowest validation loss was computationally locked down and deployed as the final model for testing.

#### 2.3.3 Model testing

Model performance was independently evaluated on the test set data by comparing model output to corresponding manual segmentation results (Figure 1-c). For a given WSI, the model was applied to sequential patches, and the results were concatenated together to generate a biopsy-level prediction at the full 40X field-of-view. Segmentation accuracy was quantified based on the dice similarity coefficient (DSC)^41^, which measures the pixel-level overlap between DL-generated segmentation results and manual segmentation results. Additionally, model accuracy, sensitivity, and specificity of sclerotic vs. non-sclerotic glomeruli was quantified based on F1, Precision, and Recall scores^42^.

#### 2.3.4 Transfer learning

The effect of transfer learning and sample size on model performance was evaluated for both non-sclerotic and sclerotic glomeruli segmentation models. Model performance metrics with and without the transferred VGG16 weights were compared and their differences quantified.

### 2.4 Deep Learning Glomerular Count vs. Standard of Care Pathology Reporting

External cases where the reported glomerular count was available (N=47) were used to compare the DL-derived glomerular count to the current standard of care. The reported glomerular counts for non-sclerotic and globally sclerotic glomeruli performed on the frozen section, along with the sclerotic-non-sclerotic glomerular ratio, were compared to both the corresponding manual segmentation-derived counts and the DL-derived counts (Figure 1-d).

Pair-wise t-tests and Pearson correlation coefficients were used to quantify statistical differences between the (a) historically reported, (b) manual segmentation-derived, and (c) DL-derived glomerular count. The Bonferroni-Holm method^43^ was implemented to correct p-values for multiple hypotheses testing. A corrected p-value lower than 0.05 was considered statistically significant.

## 3. Results

### 3.1 Manual Segmentation of Glomeruli

A total of 21146 non-sclerotic (8897 from the Internal dataset and 12249 from the External dataset) and 1322 sclerotic glomeruli (682 from the Internal dataset and 640 from the External dataset) were manually segmented on 258 images.

### 3.2 Deep Learning Detection and Segmentation Performance

DL model performance results on the External testing WSIs are summarized in Table 1. The non-sclerotic glomeruli model achieved a DSC of 0.91 (implying high spatial overlap compared to manual segmentation), an F1 score of 0.93 (implying strong overall detection performance), high recall of 0.96 (implying accurate recognition of true positive non-sclerotic glomeruli), and high precision of 0.90 (indicating a high over-prediction of non-sclerotic glomeruli compared to sclerotic glomeruli). Similarly, the sclerotic glomeruli model achieved a DSC of 0.83, an F1 score of 0.87, recall of 0.93, and a precision of 0.81. A WSI final prediction example is visualized in Figure 4.

**Table 1:** Model performance on the test set. Glomerular detection and segmentation performance for the non-sclerotic glomeruli were >0.9 by all measures, indicating robust performance. The sclerotic performance was slightly less robust but still good with measures >0.8 with recall being 0.91. The slightly worse precision compared to recall means there were more false positives than false negatives. When compared to the performance of the non-sclerotic algorithm, the precision is less robust which is consistent with the smaller amount of data in the sclerotic cohort and the relatively greater variety histologic mimics of sclerotic glomeruli on the WSIs.

|  | Detection |  |  | Segmentation |
| --- | --- | --- | --- | --- |
|  | Precision | Recall | F1 | DSC |
| **Non-sclerotic** | 0.9 | 0.96 | 0.93 | 0.91 |
| **Sclerotic** | 0.81 | 0.93 | 0.87 | 0.83 |

**Figure 4:**
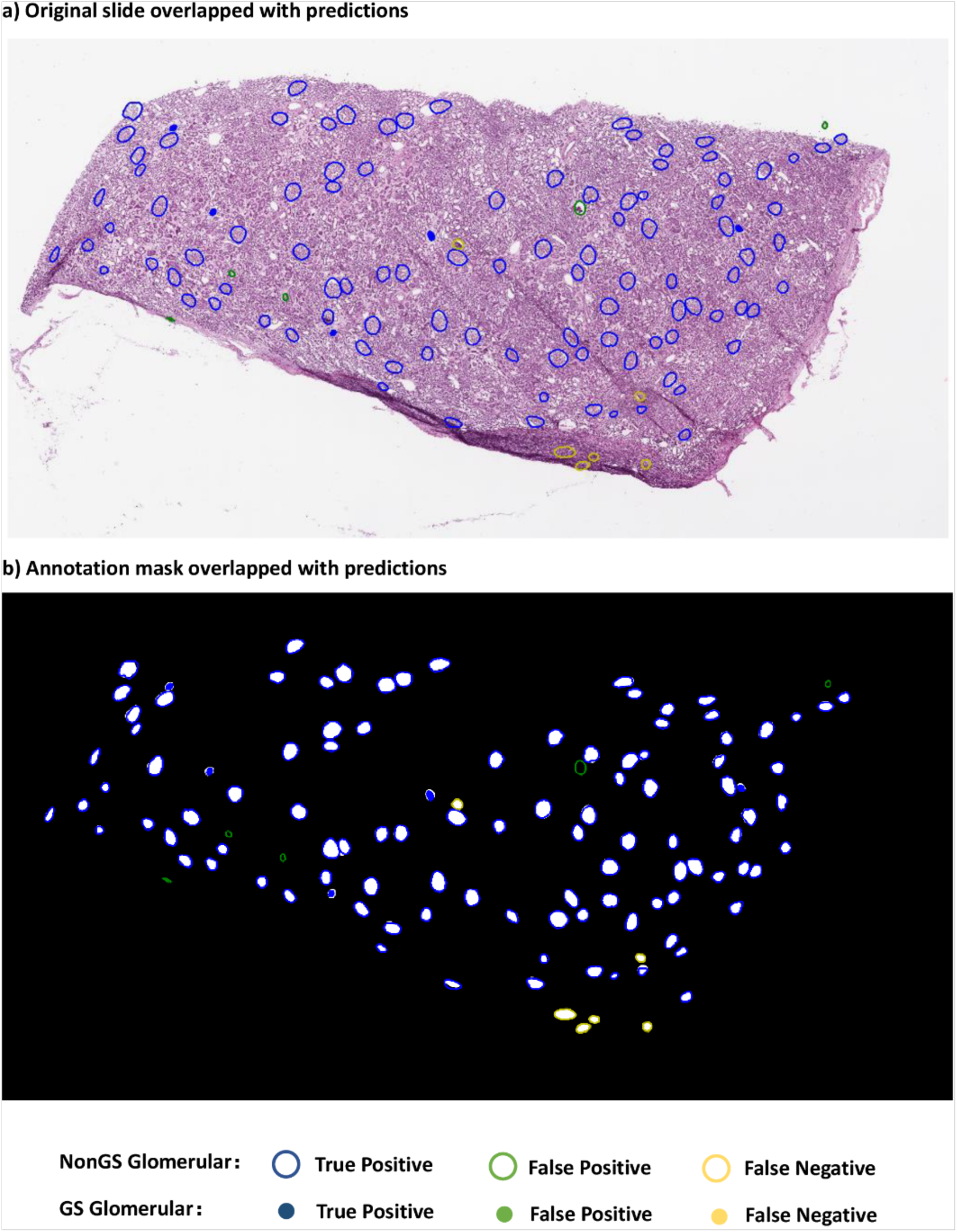
An illustrating example of a deep learning prediction on a whole slide image. Results are color coded relative to reference manual annotations to demonstrate the performance of the deep learning algorithm on testing data.

Figure 5 displays examples of false positive and false negative predictions of sclerotic and non-sclerotic models. Sources of model error included two major categories: procedure artifacts (e.g., tissue processing artifacts such as overstaining, folds, air bubbles, and chatter) and glomerular histologic mimics (e.g., fibrosis of the urinary space, dense interstitial fibrosis, red blood cell casts). The global sclerosis model had a relatively high false positive rate, due to the variety of sclerotic textures and the relative lack of global sclerosis training data. Hence, most of the incorrect predictions were due to histology mimics. The non-sclerotic model generally learned well. Extreme procedure artifacts (e.g., distorted glomeruli, tangential cuts with small glomerular profiles, and poor staining) were the major reasons for failed predictions.

**Figure 5:**
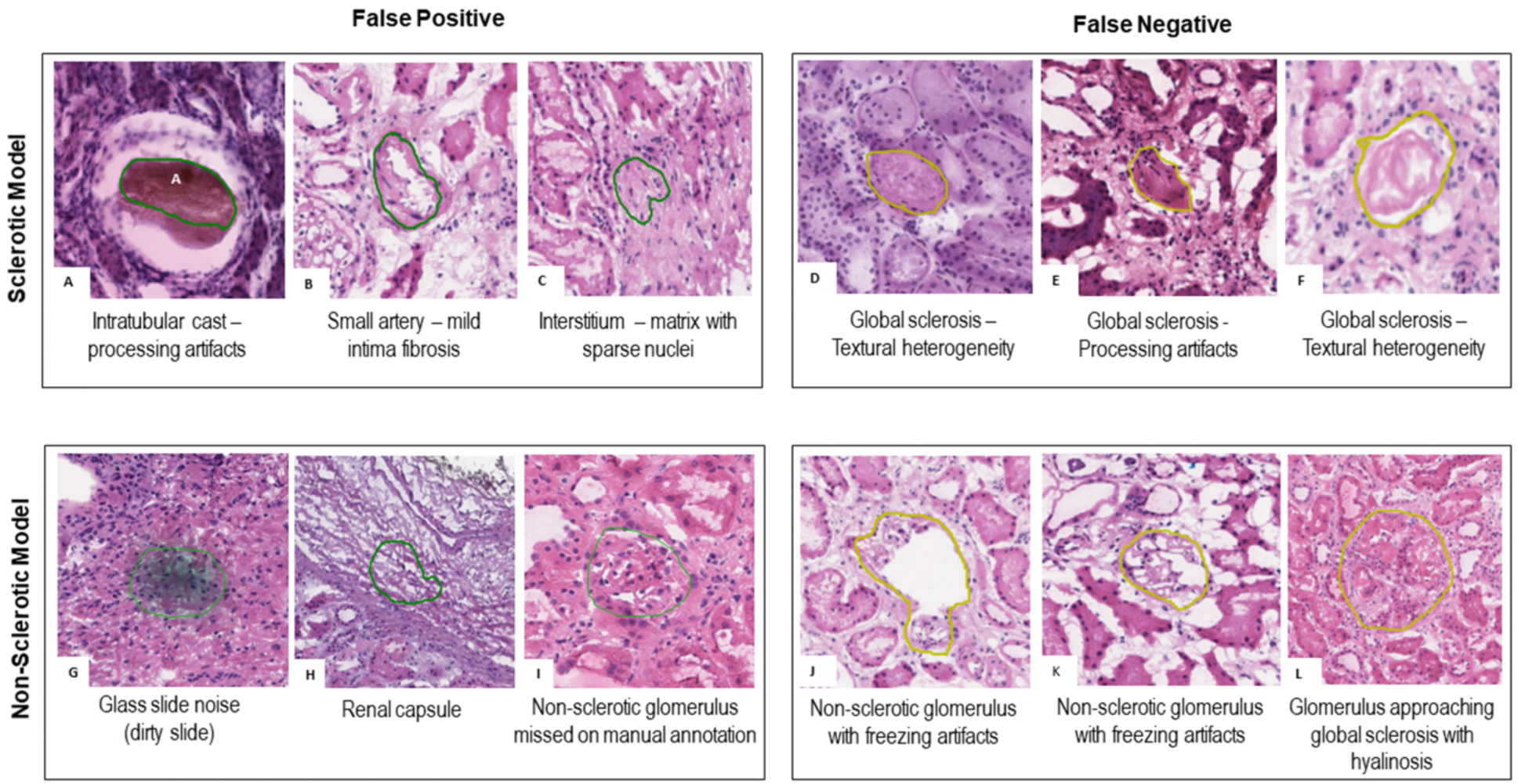
False Positive and False negative predictions for globally sclerotic and non-sclerotic glomeruli. **(A-C)** False positive for the global sclerosis model; **(D-F)** False negative for the global sclerosis model; **(G-I)** False positive for the non-global sclerosis model. A probable non-globally sclerotic glomerulus **(I)** was not manually annotated by the primary annotator nor by the quality control pathologist, but it was detected by the DL model; **(J-L)** false negative for the non-global sclerosis model.

### 3.3 Effect of Transfer Land Sample Size on Deep Learning Performance

As demonstrated in Figure 6, when training on a relatively small sample size (e.g., number of glomeruli < 600), transfer learning had a significant effect on DL model performance. Despite less data, model performance was improved based on the transfer learning procedure. The effect of transfer learning was less significant when >600 glomeruli were used for training. The non-sclerotic glomeruli model required less training samples (i.e., 1500 non-sclerotic glomeruli) to achieve performance saturation (i.e., DSC = 0.9) compared to the sclerotic glomeruli model. Meanwhile, the sclerotic glomeruli model appeared to not achieve performance saturation, implying that more sclerotic data would likely improve performance.

**Figure 6:**
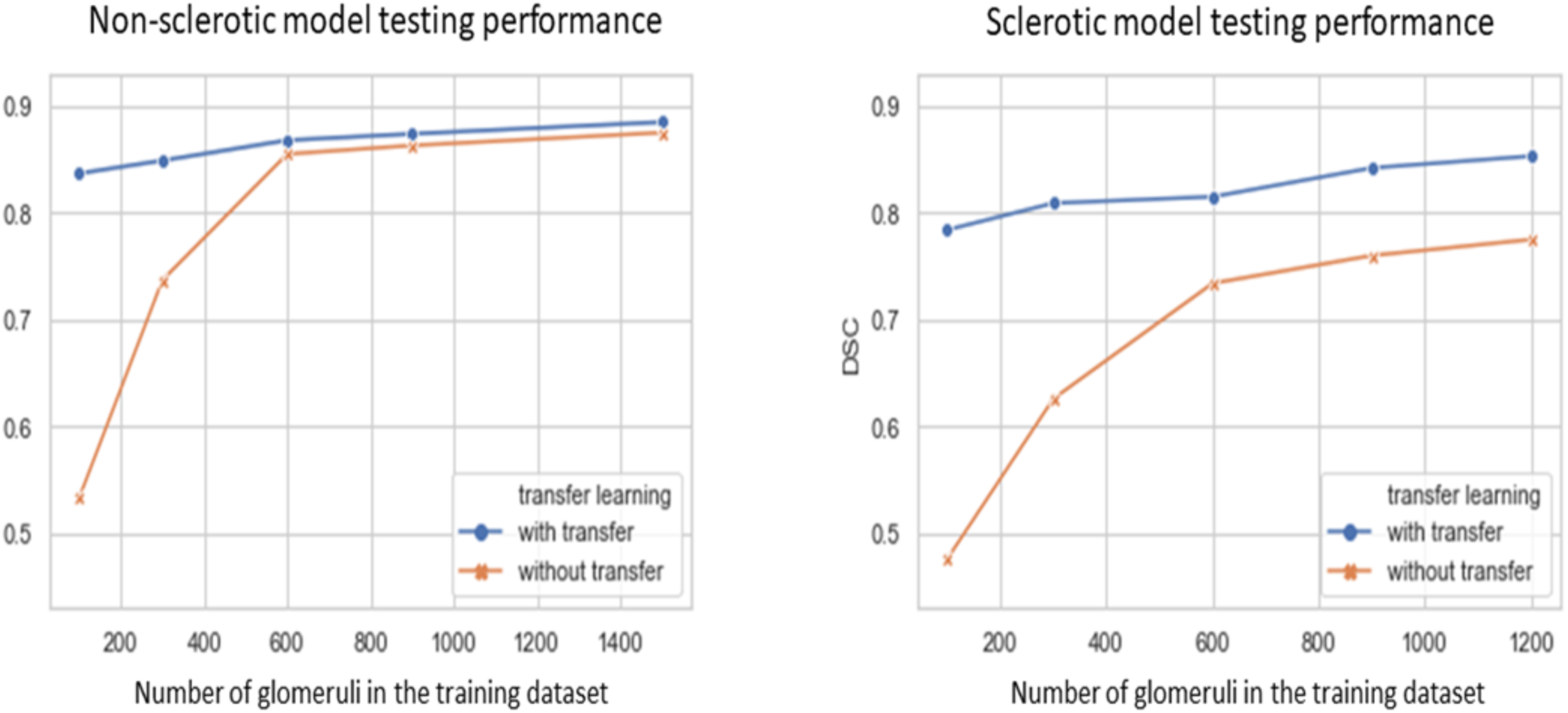
Effect of transfer learning and sample size on model performance of (left) non-sclerotic glomeruli and (right) sclerotic glomeruli. Transfer learning improved model performance significantly with limited data (i.e. <600 glomeruli) for both **(*left*)** non-sclerotic and **(*right*)** sclerotic model. Less glomeruli samples are needed for the non-sclerotic model to reach a performance saturation (i.e., DSC = 0.9 with 1500 glomeruli), compared to sclerotic model.

### 3.4 Deep Learning Glomerular Count vs. Standard of Care Pathology Reporting

Direct statistical comparisons between (i) historically reported glomerular counts, (ii) manual segmentation glomerular counts, and (iii) DL-model glomerular counts are reported in Table 2. When the glomerular count from the manual segmentation was compared to the glomerular count from the DL model, statistically similar counts were observed for both non-sclerotic (p=0.837) and sclerotic glomeruli (p=0.0950). This implies that the DL-model is operating at a non-inferior counting performance relative to an expert renal pathologist. When the glomerular counts from the manual and DL segmentation were compared to the historically reported standard-of-care glomerular counts, a statistically significant difference was observed for both non-sclerotic (p=<0.0001) and sclerotic (p=0.002) glomeruli. This implies that both manual counting by a renal pathologist and automatic counting by the DL model are both more accurate than historically reported clinical data, which is otherwise prone to high inter-observer variability. Correlation plots of the three category results among three methods for each data point are shown in Figure 7. In Figure 8, testing WSIs on the correlation plot are displayed with different procedure artifact conditions, which were a major source of model performance deviation.

**Table 2:** Comparison of glomerular count. For the model-annotation count is not significantly different from the reference segmentations, whereas the standard of care count is different from the reference segmentations. The same is true for the sclerotic model, which is not significantly different from the reference segmentations, whereas the standard of care has a significantly different count from the reference segmentations.

|  | **Model vs Reference**<br>**Segmentation** | **Report vs Reference**<br>**Segmentation** |
| --- | --- | --- |
| **Non-sclerotic** | $p=0.837$ | $p=3.695e-7$ |
| **Sclerotic** | $p=0.095$ | $p=7.920e-4$ |

**Figure 7:**
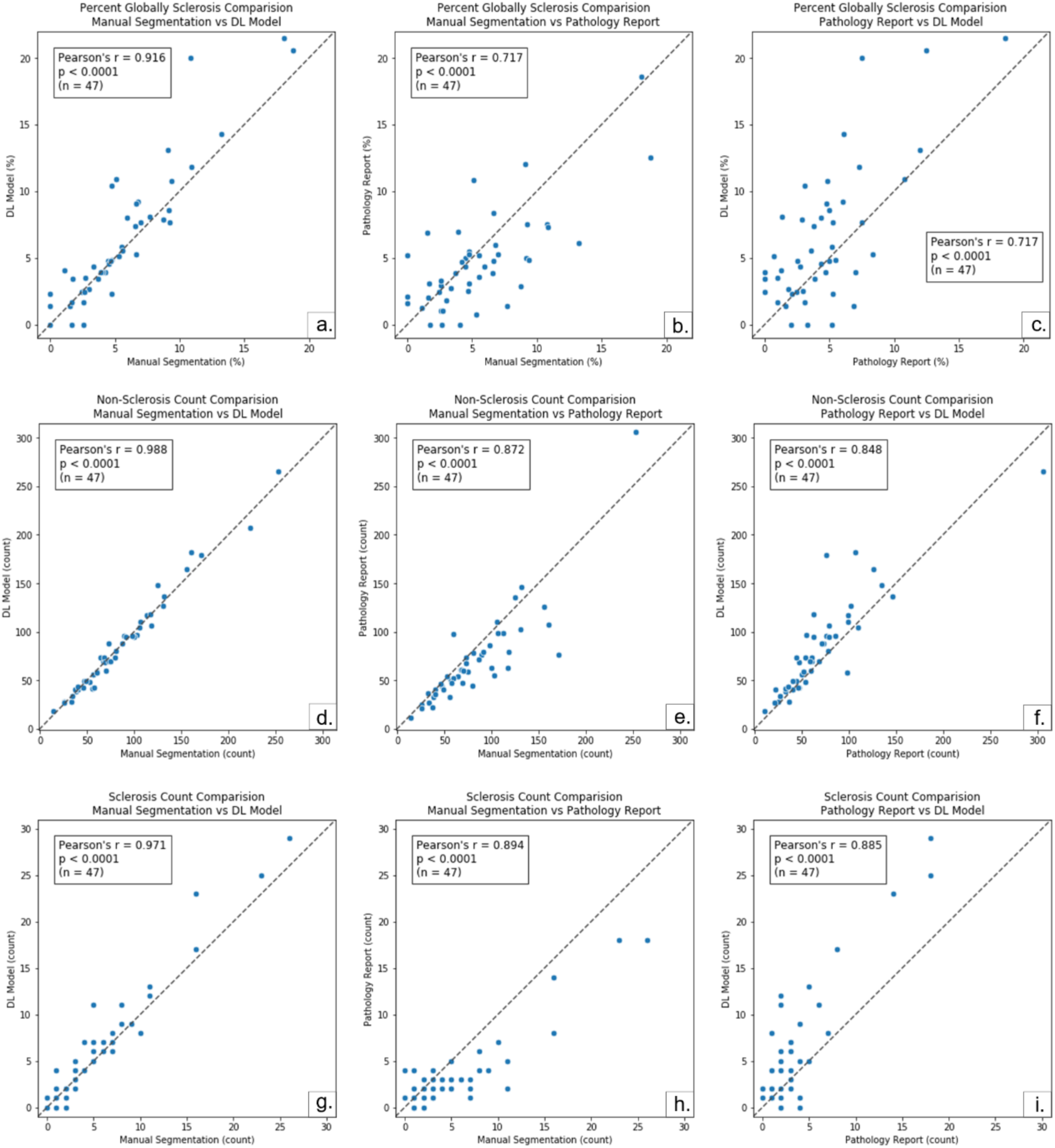
Comparison among glomerular counting modalities. Comparison for percentage of globally sclerosed glomeruli, count of non-sclerotic glomeruli, and count of sclerotic glomeruli among DL Model Prediction, Manual Segmentation, and Pathology Report.

**Figure 8:**
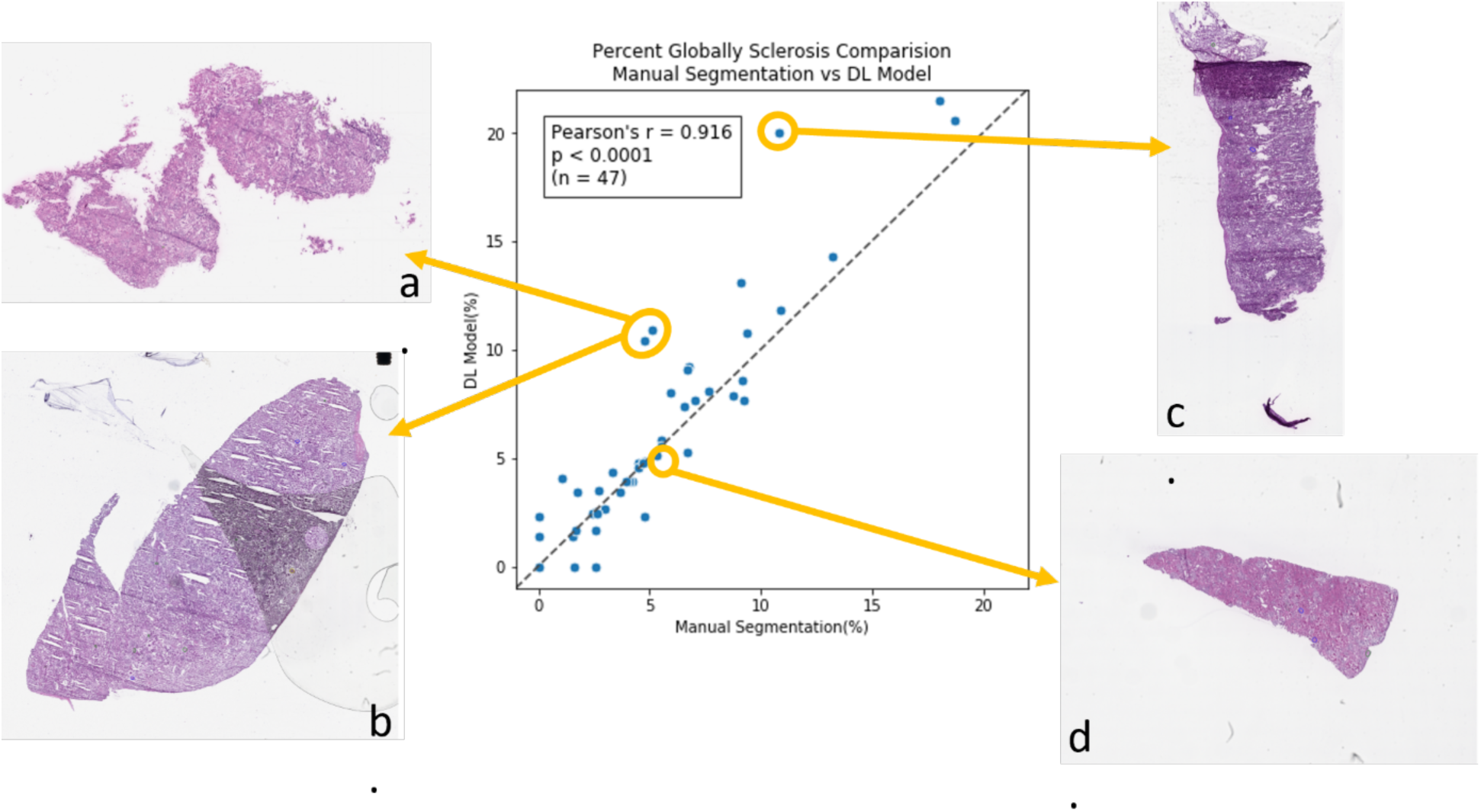
Visualization of whole slide images relative to model performance on testing data. (**a**,**b**,**c)** WSIs contain tissue freezing and folding artifacts **(a)**, glass slide (bubble between the cover slip and the glass slide) and cutting artifacts **(b)**, and tissue folding artifacts **(c)**, which may have contributed to decreased performance. **(d)** The tissue section is intact and without artifacts, with accuracy for sclerosis from the DL model prediction.

## 4. Discussion

Digital pathology and state-of-the-art computational techniques are changing the landscape of pathology practice^44^. Unlike in oncologic pathology – where computational image analysis has been extensively developed and is slowly being introduced into clinical practice, drug development, and clinical trials^45^ – native and transplant nephropathology still relies entirely on visual assessment. In the current study, we developed a robust, accurate, and generalizable DL model for automatic segmentation of non-sclerotic and globally sclerotic glomeruli on H&E frozen section WSIs from donor kidney wedge biopsies. Our results demonstrate high predictive performance on an independent dataset and show that machine-derived glomerular counts are more accurate than historically reported standard-of-care clinical data. These positive findings provide hypothesis generating data and motivate future applications of AI techniques to transplant nephropathology.

While other groups have primarily investigated DL-based glomerular detection and segmentation on WSIs of paraffin-embedded tissue^46–48^, our study represents the largest application using frozen sections from kidney donor wedge biopsies. Previous work by Marsh et. al. successfully demonstrated elliptical detection^37^ and quantification^49^ of glomeruli on frozen sections. Although their approach to DL was different than ours and their cross-validated model lacked independent testing, our key findings are comparable and thus both provide promising insight for this emerging technology.

Generalizability of a DL model is critical for future scalability in clinical practice. Even though laboratories in the United States follow general College of American Pathology (CAP)^20,34^ and Clinical Laboratory Improvement Amendments (CLIA) guidelines^35^, pre-analytical variability is still significant. Computational pathology techniques are highly sensitive to pre-analytical tissue variations^44^, including room and freezing temperature, wedge biopsy tissue thickness, the percentage of water in the biopsy tissue (freezing artifacts), frozen section thickness, the presence of folds (cutting artifacts), and composition of the stain solutions (stain artifacts)^33^. Furthermore, as cadaveric donor biopsies are obtained from deceased patients, the time from death to harvesting is often highly variable. Longer time delays from death to implantation often lead to greater autolysis artifacts on the frozen sections. Pre-existing conditions leading to patient death can also degrade tissue presentation, although these generally affect tubules and interstitium more so than glomeruli. Unlike the previously published work (37,49), our model was independently evaluated on a multi-institutional test dataset. Thus, our results provide a reasonable representation of how well the model generalized to new information not observed during training. Since independent model testing is a fundamental principle of machine learning, this is a key novelty of our research design.

While several studies used multiple pathologists to manually annotate the same WSIs to train the model on a heterogeneous group of annotations, we established a protocol where the annotation of trainees were reviewed by senior pathologists. This protocol was designed to correct the missed glomeruli due to fatigue of the primary annotator and to mimic current clinical practice, where trainees provide the first read and senior pathologists review and correct, so that a robust dataset for reference segmentations for both detection and boundaries could be generated.

Our data have also shown that the DL algorithm on digital images operates with more accuracy than clinical reads using conventional microscopy. Several reasons may account for this increased performance. First, often, pathologists assessing frozen sections donor biopsies not only operate overnight, but also are not subspecialty trained in renal pathology. Second, the spatial-visual memory of any human is limited, resulting in missing some glomeruli while counting, or counting the same glomeruli twice. The manual annotation of all the glomeruli allows for the mapping of the glomeruli on the WSIs, so that the count can be more accurate ^50^ and better serve as ground truth.

In this work, we chose to implement a UNET architecture that incorporated several state-of-the-art techniques, including transfer learning, data augmentation, and convolutional dilation ^51^. The motivation behind this design choice was based on recently published work, where UNET was successfully implemented to segment glomeruli on non-frozen tissue ^52–54^. Since our work is the first of its kind on frozen tissue, we chose a relatively simple model architecture that is commonly used in diverse biomedical image segmentation applications. We acknowledge, however, that there are newer deep learning architectures, some of which have been applied to glomeruli detection, including the Inception V3 Architecture^55^ and the SegNet architecture^56^. Future work will focus on comparing different model architectures on frozen tissue, including advanced segmentation networks such as SegNet, DeepLab V3^57^, Mask R-CNN^58^ and effective network modules such as Inception V3, Residual blocks^59^, Attention blocks^60^. While such an analysis is out of scope of the current work, characterization of different model architectures is essential to eventually implementing these new technologies in clinical practice.

Nevertheless, we believe our research design is suitable for the current dataset. First, transfer learning was shown to boost model performance by using pre-trained weights obtained from the publicly available ImageNet database as initial parameter conditions^61^. This implies that these natural images encode generalized features of quantitative image representation that are applicable to digital pathology tasks. Our results demonstrate that the relative effect of transfer learning is indirectly proportional to sample size. This finding is consistent with machine learning theory^62,63^ and implies that the performance of our sclerotic glomeruli model may asymptotically improve with more data. Second, data augmentation was also shown to increase the generalization of our DL results, suggesting artificial augmentations may capture important characteristics of renal pathology^64^. For example, *shape deformation* was shown to be effective in improving the recognition of various shapes and sizes of glomeruli. In addition, *color jitter* was useful to harmonize the large variation in stain quality, especially between the training data and the testing data. We observed sclerotic glomeruli to be most affected by *color jitter*, while non-sclerotic glomeruli were more sensitive to *texture deformation*. We hypothesize that this is due to sparser image texture in the compact structure of sclerotic glomeruli. Other augmentation methods (e.g., Gaussian blur) generally yielded better performance in the presence of freezing artifacts. Finally, our network architecture included a dilated block at the bottleneck. The rationale behind this design choice was to enable a larger receptive field-of-view to aggregate multi scale imaging information. Based on our results, we found that this dilation procedure reduced misclassification errors in the presence of tissue fold artifacts.

While our results are promising, this study has some limitations. First, all images were acquired on the same whole slide scanner. As such, it is currently unclear how scanning variability will affect the quality and performance of our trained DL models. Second, differences in model performance noted between the non-sclerotic and globally sclerotic glomeruli were due to the limited number of globally sclerotic glomeruli in the WSI dataset. We anticipate that model performance will increase based on more examples of sclerotic glomeruli. Third, our study only focused on glomeruli, which is only one aspect of pre-implant pathology. Future work will be dedicated to building additional DL models for other relevant histologic parameters such as interstitial fibrosis, acute and chronic tubular injury, vascular damage and other glomerular pathology ^65^. Additionally, our correlation analyses for glomerular counts among the DL models, the QA annotations, and the report of record were only performed on a limited number of cases where the outside reporting was available. Last, while our approach generated a robust reference segmentation dataset, inter-observer variability of segmentation was not addressed.

Digital pathology and automatic image analysis enable solutions that may aid in the clinical transplant nephropathology environment by providing robust and standardized quantitative observations, higher efficiency, centralize interpretation by expert pathologists with overall reduced error rates^66^, and by reducing the known limitations associated with visual examination ^14,17^. Additionally, a digital solution offers a more rapid and efficient allocation of the kidney overcoming the limitations, expenses, loss of precious time from the transferring of a donor organ and associated frozen section of the wedge biopsy from institution to institution, in search of a recipient.

## Data Availability

The raw data for this study can be obtained through correspondence with the corresponding
authors.

## Disclosures

### Conflicts of Interest Statement

The authors have no conflict of interests to disclose.

### Funding Statement

This work was supported by the Nephcure foundation and by Duke University institutional funding.

### Data Availability Statement

The raw data for this study can be obtained through correspondence with the corresponding authors.

## Acknowledgments

This collaborative work between the Department of Pathology, Division of AI and Computational Pathology, the Department of Medicine, Division of Nephrology, and the Woo Center for Big Data and Precision Health at Duke University.

## Notes

### Competing Interest Statement

The authors have declared no competing interest.

### Author Declarations

This study was approved by the Duke University Institutional Review Board.

